# Transmission Dynamics of COVID-19 and Impact on Public Health Policy

**DOI:** 10.1101/2020.03.29.20047035

**Authors:** B Shayak, Mohit Sharma, Richard H Rand, Awadhesh Kumar Singh, Anoop Misra

**Affiliations:** Theoretical and Applied Mechanics, Sibley School of Mechanical and Aerospace Engineering, Cornell University, Ithaca – 14853, New York State, USA; Weill Cornell Medicine, 1300 York Avenue, New York City – 10065, New York State, USA; Theoretical and Applied Mechanics, Sibley School of MAE & Department of Mathematics, Cornell University, Ithaca – 14853, New York State, USA; GD Hospital and Diabetes Institute, 139A, Lenin Sarani, Kolkata – 700013, West Bengal, India; Fortis C-DOC Hospital for Diabetes and Allied Sciences, B-16 Chirag Enclave, New Delhi – 110048, India; National Diabetes, Obesity and Cholesterol, Foundation, New Delhi, India; National Diabetes Foundation, New Delhi, India

**Keywords:** Delay differential equation, Latency and waiting time, Retardation of apparent peak, Social distancing, Effect of short-term lockdown

## Abstract

In this work we construct a mathematical model for the transmission and spread of coronavirus disease 2019 or COVID-19. Our model features delay terms to account for (*a*) the time lapse or latency period between contracting the disease and displaying symptoms, and (*b*) the time lag in testing patients for the virus due to the limited numbers of testing facilities currently available. We find that the delay introduces a significant disparity between the actual and reported time-trajectories of cases in a particular region. Specifically, the reported case histories lag the actual histories by a few days. Hence, to minimize the spread of the disease, lockdowns and similarly drastic social isolation measures need to be imposed some time before the reported figures are approaching their peak values. We then account for the social reality that lockdowns can only be of a limited duration in view of practical considerations. We find that the most effective interval for imposing such a limited-time lockdown is one where the midpoint of the lockdown period coincides with the actual peak of the spread of the disease in the absence of the lockdown. We further show that the true effectivity of imposing a lockdown may be misrepresented and grossly underestimated by the reported case trajectories in the days following the action.

## §0 INTRODUCTION

In mid-December 2019, the first cases of a hitherto unknown coronavirus were reported in Wuhan, China [1]. After a slow month and a half, the virus spread through the rest of the world with a ferocity that has taken doctors, scientists as well as public health officials by surprise. As we write this, the number of cases globally is almost 4,20,000 with more than 18,500 fatalities on record. China remains the most severely affected country with 81,000 cases, Italy is a close second with almost 70,000 and USA is third at 55,000. Whereas the rate of growth of new cases has slowed down to a crawl in China on account of the earlier start of the outbreak as well as several other factors, the growth rates in the rest of the world remain frighteningly high [2]. The worldwide explosion in the number of cases, together with a very high mortality rate in some countries, has resulted in extreme public health measures being adopted across the globe. Full lockdowns have already been imposed in Italy, Spain, France and in California and New York State in USA. These lockdowns followed a slew of increasingly strict social distancing measures such as cancellation of sporting events, suspension of classes, limitation of public gatherings to size 50, and closure of bars, discotheques and other entertainment venues. On Monday, a nationwide lockdown was declared in the United Kingdom, after weeks of deliberation and debate on the adverse socio-economic impacts of such a move, as the growth in new Coronavirus cases kept accelerating relentlessly. Just yesterday, a similar move was announced in India, and today in New Zealand, both ostensibly less-affected countries, after several days of steady and continuous increments in the numbers of cases.

### ETIOLOGY AND TRANSMISSION CYCLE OF THE PATHOGEN

Coronavirus (CoV) belongs to *Coronaviridae* family of order *Nidovirale*. CoV are single-stranded RNA viruses (+ssRNA) having a spike glycoprotein on the envelope, giving it a crown-like appearance when seen through an electron microscope. The subfamily *Orthocoronavirinae* has four genera of CoVs: Alpha-coronavirus, Beta-coronavirus, Delta-coronavirus and Gamma-coronavirus. Furthermore, the genus Beta-coronavirus is divided into five sub-genera or lineages. While bats and rodents are considered to be the genetic sources of alpha and beta-coronavirus, avian species represent the genetic sources of delta and gamma-coronavirus [3].

To date, seven human coronaviruses (HCov) have been identified. The A lineage of beta-coronavirus (such as HCoV-OC43 and HCoV-HKU1) as well as alpha-coronavirus (such as HCoV-229E and HCoV-NL63), causes common colds and self-limiting respiratory infections. In contrast, the B lineage of beta-coronavirus includes SARS-CoV (severe acute respiratory syndrome) and SARS-CoV-2 (the pathogen behind COVID-19), while the C lineage of beta-coronavirus includes MERS-CoV (Middle East respiratory syndrome). Both the latter lineages are responsible for recent epidemics with a variable clinical severity of pulmonary and extra-pulmonary involvement and associated significant morbidity and mortality [4].

SARS-CoV-2 is a single-stranded, positive sense RNA virus, having a diameter of 60–140 nm with a round or elliptic shape; however, it often exists in pleomorphic form. Its RNA genome contains 29891 nucleotides, encoding for 9860 amino acids and shares 99.9% sequence identity, suggesting a very recent host shift into humans [5,6]. Like other CoVs, it is sensitive to ultraviolet rays and heat. Besides, these viruses can be effectively inactivated by lipid solvents including chloroform, ether (75%), ethanol, peroxyacetic acid and chlorine-containing disinfectant but not by chlorhexidine [5,6].

Coronaviruses are naturally hosted by bats and it is believed that most human coronaviruses are derived from the bat reservoir [7,8]. Genomic sequence studies of COVID-19 have identified nearly 50%, 79% and 96% similarity to MERS-CoV, SARS-CoV and bat SARS-related coronavirus respectively [9,10,11]. The specific route of transmission to human from natural reservoirs is still yet to be known, however, some studies suggest that pangolins could be the intermediate mammalian hosts. Since the spike proteins of SARS-CoV-2 are nearly identical to those of a virus isolated from pangolin, it is believed that pangolins could have provided a partial spike gene to SARS-CoV-2, to infect mammals [12,13]. A recent study has also shown a development of new variations at the functional sites in the receptor-binding domain of the spike of SARS-CoV-2 and viruses from pangolin, likely caused either due to a natural selection or due to mutations and recombination, or both [14].

Nevertheless, once a human is infected, the virus can be transmitted from human-to-human through the respiratory droplets and aerosols from coughing and sneezing. This is similar to the transmission mechanism of other respiratory pathogens, including SARS-CoV. SARS-CoV-2 also uses the angiotensin converting enzyme II (ACE2) receptors like the SARS-CoV [15].

The incubation period of COVID-19 can vary from 3 days to 14 days with a median of approximately 5 days [16]. The epidemic in Wuhan, China also doubled about every seven days during the initial phases. On average, each patient transmits the infection to an additional 2.2 individuals, suggesting a basic reproduction number *R*_0_ of approximately 2.2.

### RELATION TO PAST EPIDEMICS

Here we focus on the relationship between COVID-19 and the past outbreaks of SARS, MERS and swine flu (H1N1). Epidemics of infectious diseases have disastrous impacts on health, social and economic conditions in developing as well as developed countries. To control the spread of these diseases, it is crucial to effectively utilize public health resources in a timely manner. The process of anticipating, preventing, preparing for, detecting, responding, and controlling epidemics in order that the health and economic impacts to society are minimized is described as epidemic management. A well-coordinated, systematic and expeditious investigation by government strategists, policy makers, physicians and technical staff is required to understand the outbreaks and identify key points for implementing interventions. Such interventions include adopting hygienic measures, social distancing, travel restrictions, quarantine and case isolation. We shall now discuss the applications of these methods to the SARS, MERS and Swine Flu outbreaks.

The SARS outbreak started in Guangdong, China in 2002 and spread across 29 countries, causing 8096 cases and 774 deaths, for a case fatality rate (CFR) of 9.6 percent. SARS is primarily transmitted by respiratory droplets, direct contact and fomite-based contact and has an *R*_0_ of approximately 2.5-3.0 [17-19]. During the outbreak, people with suspected illness were isolated from the hospitals, those travelling from SARS affected countries were instructed to monitor their health for 10 days and visit a hospital in case of any symptoms [20], and close contacts of reported cases were quarantined. A study [21] demonstrated that always wearing a mask when going out was associated with 70% reduction in risk compared to never wearing a mask. A second study conducted in Taipei [22] showed that having checkpoint alcohol dispensers for glove-on hand rubbing between zones of risk, along with thermal (fever) screening at special stations outside the emergency department, helped in effectively minimizing nosocomial SARS infection of HCWs.

MERS was first reported in Saudi Arabia (the country lacking the requisite geopolitical clout to detach the name of the originating region from the name of the disease) in 2012, and spread to 27 countries, where it was responsible for 2494 confirmed cases and 858 deaths, amounting to a staggeringly high CFR of 34.4 percent. Public health management of MERS involved infection control strategies for hospitalized patients and healthcare professionals. Hand hygiene, careful use of personal protective equipment, patient isolation and the use of engineering controls in hospitals were some of the measures adopted for preventing the spread of this deadly disease.

In 2009 there was an outbreak of swine flu (H1N1) or Influenza type A virus, in the United States, which affected 1.4 billion people worldwide within a year and caused more than 18000 deaths. CDC believed that this virus was a result of reassortment, a process through which two or more influenza viruses exchange genetic information by infecting a single human or animal host. A public health emergency was declared back then, but the death rate was lower than that of the seasonal flu (between 0.001 and 0.007 percent). This prevented the epidemic from escalating into COVID-19 proportions. Further, several vaccines were later developed which has significantly brought down the death rate since then.

The pathogen behind the current pandemic COVID-19 derives its uniqueness from its molecular structure, discussed above. Even casual contact such as handshakes is sufficient to spread this deadly virus, and its epidemiology makes it hard to detect this infection at an early stage. Scientists have found that the pathogen remains on surfaces for a long time ranging from 3 hours in aerosols, 24 hours on cardboard and up to 3 days on plastic and stainless steel. Worse still, a large number of affected patients don’t show symptoms for five or more days after exposure.

When developing a mathematical model for COVID-19, we must be able to use it to explain why COVID-19 has assumed the terrifying proportions which SARS and MERS did not. Before presenting our model, let us take a brief look at the methods already in existence for modelling epidemic diseases. In the rest, residue and remainder of this Article, we shall use the standalone term “Coronavirus” to denote the pathogen SARS-CoV-2 responsible for COVID-19; the other coronaviruses will always be referred to explicitly.

### MATHEMATICAL MODELING TECHNIQUES

The first attempt to model the spread of an epidemic though a set of differential equations was by WILLIAM KERMACK and ANDERSON MCKENDRICK in 1927 [23] who constructed a third order nonlinear model. Their basic variables were *x*, the number of people susceptible to the epidemic, *y* the number of people infected by it and *z*, the number of people recovered from it (or killed by it). Their pioneering study formed the basis of the S-I-R (susceptible-infected-recovered) model which has since been applied to influenza [24], dengue [25] and Ebola virus outbreaks [26]. Numerous modifications to the basic model have been proposed over the years; the one which is most relevant for us is the addition of delay [27,28]. In these works, the delay represents the incubation period i.e. the gap between a person’s being exposed to the pathogen and his/her turning infectious. These and similar models may be considered as lumped-parameter models, in the sense that they feature a small number of parameters which represent averages over various kinds of population, for example the young and the old, the healthy and the constitutionally infirm. The advantage of these models is that they are considerably simple, easy to use, and yet capable of yielding the broad trajectory of the epidemic in a region with high accuracy.

Dynamic modeling of COVID-19 along these lines has been attempted in several prior works [29-34]. Kucharski et. al. [29] modelled the early phases of the outbreak in Wuhan to estimate the value of *R*_0_ there. The number they obtained was between 1.6 and 2.6. A similar study, determination of *R*_0_ as a function of time, has been performed by Feng et. al. [30], whereas an extensive determination of parameters in the S-I-R model has been done by Rabajante [31]. Peng et. al. [32] use a similar model but with a different purpose – they predict the long-term course of the outbreak in Wuhan as well as in the rest of China. Chen et. al. [33] have used a more complicated model with 14 dynamical variables to explain the transmission of the virus from bats to humans and thereafter its spread in the human population. Neher et. al. [34] have introduced a parametric excitation into the basic S-I-R model to study the effects of seasonal temperature variation on the spread of the epidemic. As alternative approaches to the modeling, we would like to cite References [35,36] which have used stochastic models to analyse the course of the disease in Wuhan and on the cruise ship Diamond Princess respectively. A mathematical analysis of the consequences of public health policy is Reference [37] which predicts the effects of various kinds of social distancing measures such as school closures and social isolation of the high-risk senior citizens. The models in this paper are based on individual-based stochastic influenza models [38]. As recently as the day before yesterday, a new study [39] has emerged which explores the effects of non-pharmaceutical interventions (social distancing etc) on the spread of COVID-19 in Singapore. The mathematical core of this work is a stochastic model, once again for influenza [40]. Another recent work [41] attempts to find the required hospital capacity in Chicago in the presence and absence of a lockdown, using the model [34]. Yet another contribution [42] predicts the effects of imposing a lockdown in India, using an in-house lumped-parameter model. This model is based on S-I-R and accounts for the variation in susceptibility across different age groups. It also accounts for quarantining.

One of the features indigenous to this new and dangerous Coronavirus strain is that there are time lags involved in several stages of the transmission dynamics. One major delay step is that patients become transmissible before showing symptoms of the disease [43]; in other words, there is a time gap or **latency period** between the time a patient turns infectious and time that s/he manifests symptoms. A second delay step comes from the fact that, on account of the element of surprise associated with the pandemic, testing facilities in many countries and regions are inadequate relative to the number of cases taking place. For example, the testing rate in USA increased by a factor of ten from 03 March to 17 March [44], with some states such as New York achieving the maximum testing rates. Similarly, in India, Russia and some European countries, the testing rate is also undergoing a sharp increase even as we write. A limitation in the number of testing facilities means that there is a waiting period involved between the time that a patient reports with symptoms to an isolation-cum-testing facility and the time that the test results actually arrive. In South Korea on the other hand, there was a huge number of testing centres to begin with and this wait was very small. The role of testing delay in the Ebola epidemic has been discussed in Reference [45].

In Section 1 of this work, we construct a lumped-parameter dynamic model of the transmission of COVID-19 which accounts for this novel feature of the Coronavirus. In Section 2, we validate the model using test cases. In Section 3 we use the model to predict the effects of the delays as well as to analyse the consequences of imposing harsh social restrictions such as lockdowns for a limited period of time. We wrap things up with a Conclusion.

## §1 CORONAVIRUS DYNAMIC MODEL

We focus on the time evolution of the epidemic in one localized region, for example a city, district or county, which has sufficient inter-mixing of its population but zero or limited interaction with outside locations. Our basic model is derived from traditional S-I-R models but we have made some important changes to account for the unique nature of this disease. Specifically, we must be able to account for the following features :

- Quarantine of patients as well as anticipatory quarantines of potential patients are very important parts of the epidemic management process.
- Even symptomatic patients carry significant viral load prior to manifestation of symptoms. Thus, they can transmit the disease to other people before being quarantined or otherwise isolated from society.
- There are asymptomatic carriers of the disease who are not quarantined or isolated except to the extent required by society and/or law.
- Due to limited testing facilities, there can be a significant waiting time between a patient arriving for testing and the results being received.

To account for these features, we define four basic variables which are somewhat different from the conventionally chosen variable set. Before introducing them, we state a definition. A **sick** person is defined as one who gets infected with the Coronavirus at some stage of the epidemic’s evolution, at such a level as to make him/her capable of transmitting the virus to others. The person becomes sick as soon as the virus reaches a transmissible level in his/her system. It does not matter whether the case is asymptomatic or symptomatic. Once sick, the person’s status remains sick for all time, even after recovering from the disease. The assumption underlying this definition is that the same patient cannot be infected more than once during the course of the epidemic. This is a very reasonable assumption on account of the immunity arising from a past infection. A **healthy** person is one who is not sick. Note that for the purposes of this study, people suffering from other diseases, especially flu, are classified as healthy even if they be in a critical condition.

Given this, our four variables are

- *x* (*t*) : the number, at any given time, of susceptible but healthy people at large i.e. not in quarantine or enforced total isolation. By susceptible, we exclude that fraction of the healthy population who are immune to the virus or in whose system the virus cannot take hold.
- *y* (*t*) : the number, at any given time, of sick people who have had some exposure to society. This includes asymptomatic i.e. undetected cases as well as people who later report for quarantine and testing after developing symptoms. It excludes people who are already in anticipatory quarantine as a result of contact tracing, and fall sick there.
- *z* (*t*) : the number, at any given time, of people who are in quarantine or enforced isolation and are scheduled for testing.
- *w* (*t*) : the number, at any given time, of people who have tested positive for the virus.

The reason behind our variable choice is that *y* is an excellent indicator of the actual spread of the disease in free society (as opposed to in anticipatory quarantine) while *w* is the time history of cases which gets reported in official records of the disease. A disparity between these two is something we wish to highlight. Finally, we note that like all lumped-parameter epidemic dynamic models, ours is a good one for the bulk of the disease but is not expected to work well during the very early stages of spread and the final phases of trail-off.

We shall now begin the derivation of the Coronavirus dynamic model. The derivation is involved, and the final model itself is (7). The flow diagram of people from one category to the other looks like the following, where *R* denotes a flow rate (i.e. d/d*t* of a population).

In this schematic, *R*_1_ is the rate at which healthy, at-large people fall sick, *R*_2_ is the rate at which people arrive for quarantine and testing, *R*_3_ is the rate at which positive results are detected and *R*_4_ is the rate at which negative results are detected. We take for granted that every person who is tested for the virus is in quarantine. We shall now elaborate on the functional forms of these rates.

Tradition has it to make *R*_1_ proportional to the number of healthy (and susceptible – even when we don’t mention this explicitly) people and the number of sick people, i.e. to make *R*_1_ have the form *kxy* where *k* is some constant. We follow the tradition with a variation. First, recall that by our definition, sick people stay sick for all time. However, they do not transmit the disease for all time. Consider an asymptomatic carrier who remains exposed to society throughout. We assume that the number of asymptomatics is proportional to the total number of sick people exposed to society, i.e. we let the number of asymptomatics be *μ*_1_*y* where *μ*_1_ is a number between 0 and 1. After falling sick, an asymptomatic carrier can transmit the disease for 7-10 days, which is called the infection period [46]. Thus, new healthy people can be only be infected by those asymptomatic sick people who have fallen sick within the last 7-10 days, and not those who have fallen sick earlier. The number of such people is the number of asymptomatic sick people today minus the number of those 7-10 days earlier i.e. it is *μ*_1_ (*y*(*t*) − *y*(*t* − *τ*_1_)) where *τ*_1_ is the infection period. This *τ*_1_ has the structure of a delay term, although it is not one of the fundamental delays mentioned in the Abstract and Introduction. Thus, we get a contribution to *R*_1_ of the form *k*_0_*xμ*_1_ (*y*(*t*) − *y*(*t* − *τ*_1_)) where *k*_0_ is the basic proportionality constant, and one of the most fusndamental parameters in our model. Its value depends on the reproductive number *R*_0_ of the virus and on the degree of interaction prevalent in the society – the greater the social distancing, the smaller the value of *k*_0_. A constant value of *τ*_1_ represents an averaging process over various populations. There is a second, and equally dangerous, contribution to *R*_1_. This comes from the sick people who will eventually turn symptomatic and isolate themselves but are still transmissible, blissfully unaware of the fact. These account for 1−*μ*_1_ of the total exposed sick people, and they transmit for a smaller duration, say 2-3 days, before developing symptoms and reporting for quarantine. This duration, which we call *τ*_2_, is the latency period. We can repeat the preceding argument to obtain the contribution of these people to *R*_1_ as *k*_0_ *x* (1 − *μ*_1_)(*y*(*t*) − *y*(*t* − *τ*_2_)) where the effective *τ*_2_ is once again an average over various populations. Putting these together we have

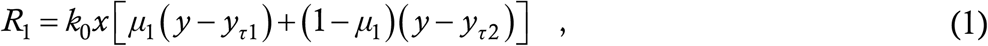

where the notation *y*_*τ*1_ means *y* delayed by time *τ*_1_, *y*_*τ*2_ means *y* delayed by *τ*_2_ etc. The absence of *z* and *w* in *R*_1_ incorporates the assumption that there is no violation of quarantine by symptomatic patients and that recovered patients are kept in quarantine until they are no longer transmissible. The effects of relaxing these assumptions are being reserved for a future study.

Turning now to *R*_2_, we have already indicated its various components in Fig. 1. The first component, *R*_2*a*_, requires the most thought to express in a mathematical form. By hypothesis, at this moment, during an infinitesimal time interval Δ*t, R*_1_Δ*t* number of people take sick, of whom *μ*_1_*R*_1_Δ*t* will be asymptomatic (zero contribution to *R*_2*a*_) and (1−*μ*_1_)*R*_1_Δ*t* will be symptomatic (full contribution to *R*_2*a*_). But, this latter fraction will manifest symptoms and hence get to know of their sickness during a time interval Δ*t* **at a time *τ***_**2**_ **into the future from now** and head to quarantine only then. Thus we can say that, at time *τ*_2_ from now, the rate of progress to quarantine will be (1−*μ*_1_)*R*_1_ where *R*_1_ is measured now. In other words,

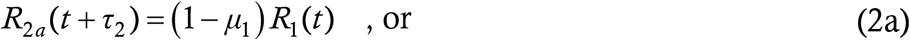

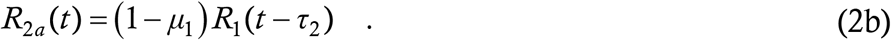

**Figure 1:**
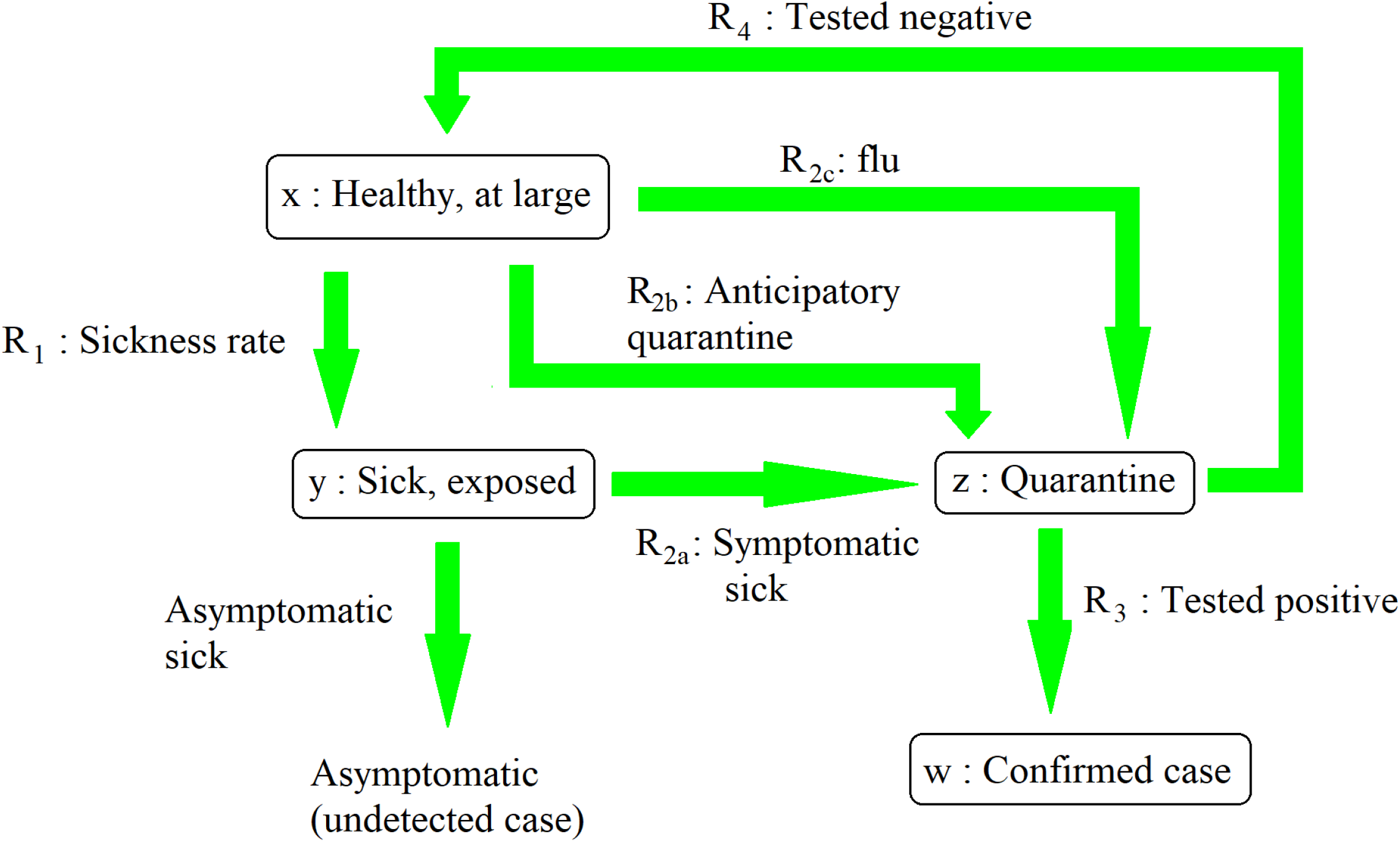
Schematic diagram showing the flow of people to and from the various categories, and defining the rates R_1_ to R_4_

Thus, the time lag or latency period between falling sick and appearance of symptoms adds a delay to the rate. In other words, the flow rate out of exposed sick is (the relevant fraction of) the flow rate in, **retarded by the time *τ***_**2**_. The logic leading to the retardation is important – we shall use it again. The other contributions to *R*_2_ are easier. *R*_2*b*_ is the number of anticipatory quarantines – these are typically made by contact tracing of existing cases, so it is reasonable to assume a form *k*_3_*xw*. This assumes that the tracing and quarantining are done nearly instantaneously, which is reasonable since the time scale involved is usually a few hours and not days. The proportionality to *x* in this term ensures that if the number of healthy people at large is very small, then also huge numbers don’t get quarantined through contact tracing. Finally, *R*_2*c*_ represents the fact that there are some people who do not have the Coronavirus but display similar symptoms as a result of other illnesses for example flu. These people also present to quarantine and testing. We model this through a term *k*_4_*x*. Adding all these together,

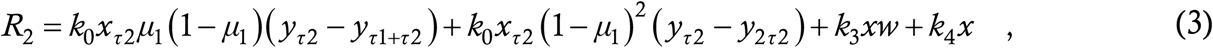

where the subscripts on *x, y, z* and *w* indicate the times through which they are retarded.

*R*_3_, the flow rate from quarantine to positive, consists of all the people who entered quarantine through *R*_2*a*_ plus a certain fraction, call it *μ*_2_, of people who entered anticipatory quarantine as a result of contact tracing. Due to the limited number of testing apparatus and huge queues for these facilities, it is often the case that several days elapse between a patient being admitted and his/her test results being obtained. Thus, quarantine is again a holding stage like exposed sick, with the delay here being some interval *τ*_3_. Following the logic of the last paragraph, all rates out of quarantine will be retarded by *τ*_3_ just as the rate *R*_2*a*_ out of exposed sick was retarded by *τ*_2_. Hence *R*_3_ is *R*_2*a*_ retarded by *τ*_3_ plus *μ*_2_*k*_3_*xw* again retarded by *τ*_3_ i.e.

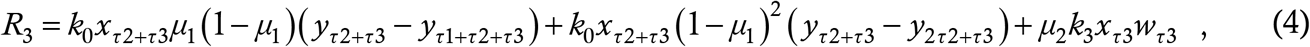

where we can see various combination delays now entering the picture.

Finally, *R*_4_ consists of the fraction 1−*μ*_2_ of anticipatory quarantines plus all the flu cases, the whole retarded by *τ*_3_ i.e.

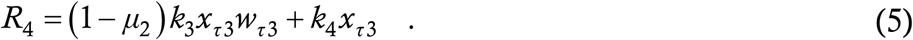

This completes the determination of the various rate equations.

The flow diagram (Fig. 1) tells us that the time evolutions of the variables are (using overhead dot for time derivative as is customary)

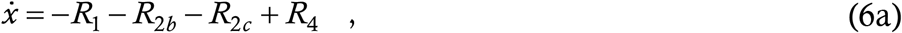

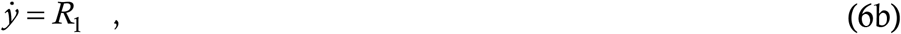

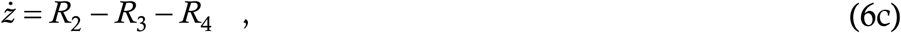

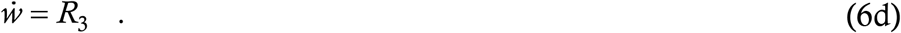

Note that the d/d*t* of *x* + *y* + *z* + *w* is not zero but equals *R*_2*a*_, since some people who are in *y* again show up in *w*. Hence *x* + *y* + *z* + *w* double-counts part of the population and has a positive time derivative, as is consistent (we have explained earlier the advantage of our variable choice). Finally, we have not treated the number of deaths as a separate variable. The tacit assumption is that the mortality count is a fixed percentage [47] of the number of symptomatic cases *w*, with the percentage depending on various factors such as average age of the population and quality of healthcare services. From our viewpoint, a dead person is as capable of transmitting disease as a person recovered in quarantine, so *w* can take care of both of them.

Combining the six previous equations leads to the final form of the Coronavirus dynamic model :

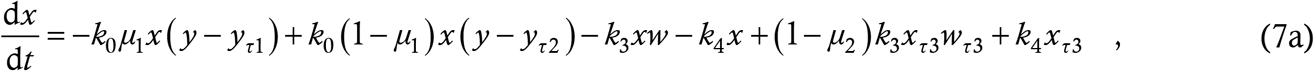

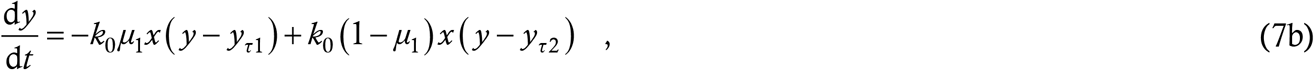

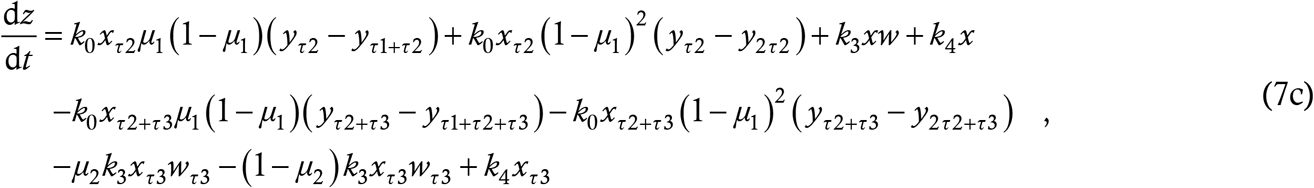

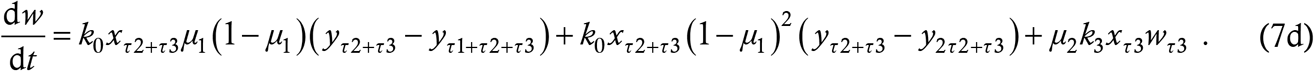

Just to recapitulate for those who have skipped the derivation, the parameters here are *k*_0_ : the basic transmission rate of the disease which includes factors such as *R*_0_ of the pathogen and degree of social distancing, *k*_3_ : a proportionality constant expressing the number of anticipatory quarantines, *k*_4_ : a proportionality constant reflecting the number of flu cases, *μ*_1_ : the fraction of asymptomatic carriers, *μ*_2_ : the fraction of anticipatory quarantines who test positive, and the three delays *τ*_1_ : the duration that an asymptomatic carrier is transmissible, *τ*_2_ : the latency period i.e. the interval between start of transmissibility and manifestation of symptoms and *τ*_3_ : the interval between reporting for testing and the results being obtained. This is a huge number of parameters – we shall see later that the most important are *k*_0_, *τ*_2_ and *τ*_3_.

## §2 SOLUTION OF THE MODEL, TEST CASES

The dynamic model (7) is impossible to solve analytically so we use numerical integration. The method we have opted for is 2^nd^ order Runge-Kutta similar to the routine dde23 [48] in Matlab. We did not use the Matlab routine however because of the huge number of delays involved in this problem; instead we wrote our own solver. We tested the solver against the results of a previous paper [49] where we had used dde23 along with various theoretical methods. Since all results of that paper were reproduced, we believe that the solver is accurate.

We shall now solve the model for some test cases to demonstrate its plausibility. We measure time in days, from *t* = 0 to *t* = 100, at which point we assume that the epidemic is over (for example through change in temperature, mutation of the virus, development of a cure etc). We use the time step *h* = 0.001 day. We assume that the population of the region is 500 units (for the plots we shall normalize to unity but for the simulation, a larger number proved convenient). The **benchmark parameter values** we consider are *k*_0_ = *k*_3_ = 0.0008, *k*_4_ = 0.0004, *μ*_1_ = 0.18, *μ*_2_ = 0.02, and the delays *τ*_1_ = 7, *τ*_2_ = 3 and *τ*_3_ = 5 (we shall explain some of the choices later). A delay differential equation needs to be seeded with initial functions lasting as long as the maximum delay involved in the problem. Here, this maximum is 15. So we have gone with the initial functions *x* (*t*) = 500 − *t*/15, *y* (*t*) = *t*/15, *z* (*t*) = 0 and *w* (*t*) = 2 × 10^−6^ for the time interval [0, 15]. This assumes a slow growth of the epidemic during the seeding period. The time traces of *x, y, z* and *w* for this run are shown in the Figure below. Here and henceforth, we plot *x* in blue, *y* in green, *z* in red and *w* in grey. In the rate plots which appear later, we shall use the same colour scheme for the time derivatives of these four variables. We also normalize the initial population to unity.

The results are physically plausible with the number of healthy at large people (blue) decreasing over time and the various other populations increasing over time. Before proceeding further, we briefly comment on the parameter values for this run. The values of *τ*_1_, *τ*_2_ and *μ*_1_ come from virological studies and are approximately independent of region. For *τ*_2_, Reference [43] reports that patients admitted to hospital with COVID-19 already carried significant viral loads. Since the median incubation period for COVID-19 estimated there and elsewhere [16] is about 5 days, we have taken *τ*_2_ to be 3 days, or approximately half this period. The benchmark *μ*_1_ comes from Reference [50]. The parameters *τ*_3_, *k*_3_, *k*_4_ and *μ*_2_ on the other hand are region-specific and they come from records collected at the testing facilities in the region. Here we have gone with average, plausible values – when modeling a specific region they will need to be determined from the data. The most important parameter in the model is *k*_0_ which has both virological and region-dependent contributions – here we have chosen it so as to generate a peak in reported cases at about 35-40 days after initiation. We have found that the results which we shall claim in the subsequent pages are to a large degree independent of the exact numerical values of the parameters and hold true whenever there is a general spreading profile of a sharp peak surrounded by flats on both sides. Later in this Section, we shall give an example of how to estimate the parameters by fitting the model to an available data set. We note that for the benchmark run, the fraction of healthy at large people remaining at the end of the 100-day period is 31 percent of the initial value. This number will be useful in comparing the disease trajectories as the parameters are varied.

We now run some test cases, where we intend to compare COVID-19 with SARS and MERS. The latter comes first. MERS virus has an *R*_0_ of just above unity [51] as against 2.2 or more for the Coronavirus. In our model, reproduction number and hence transmissibility is taken into account through the factor *k*_0_. Accordingly we reduce *k*_0_ from 0.008 to 0.006 and plot the run below.

There is a drastic increase in the ultimate number of healthy at large people, to more than 80 percent of the initial population. In a series of test runs (not shown here), we also find that increasing the value of *k*_0_ causes the peak in the transmission rate to appear earlier and be higher, which is consistent with the known fact that social distancing broadens and flattens the peak. The maximum value of *z* (number of people in quarantine) also increases significantly with increasing value of *k*_0_.

We now bring SARS into the picture. The *R*_0_ for this virus was very high, at about 2.5-3 [19]. However, once again the epidemic did not escalate to the level of COVID-19. This can be ascribed to the fact that in SARS, the affected patients manifested symptoms before becoming significantly transmissible and thus could be quarantined early. On the other hand, as we have mentioned before, COVID-19 has a latency period of *τ*_2_ between the beginning of transmissibility and the appearance of symptoms. To test the sensitivity of the epidemic trajectories to this latency period, we repeat the simulation with the parameters of Fig. 2 but with *τ*_2_ reduced from 3 to 1.5. The results are presented below.

**Figure 2:**
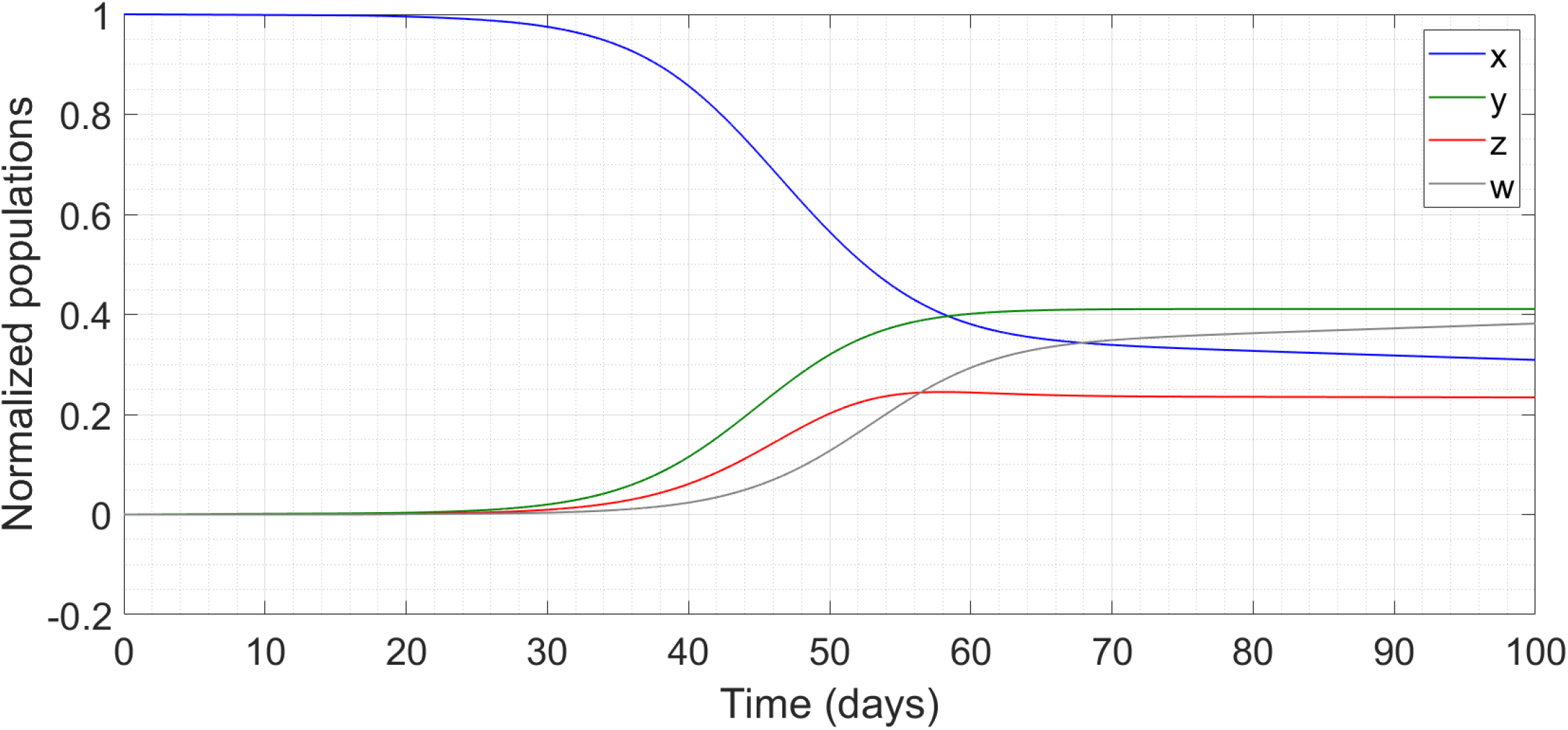
Simulation of the benchmark model, showing time traces of x, y, z and w during the course of the epidemic.

**Figure 3:**
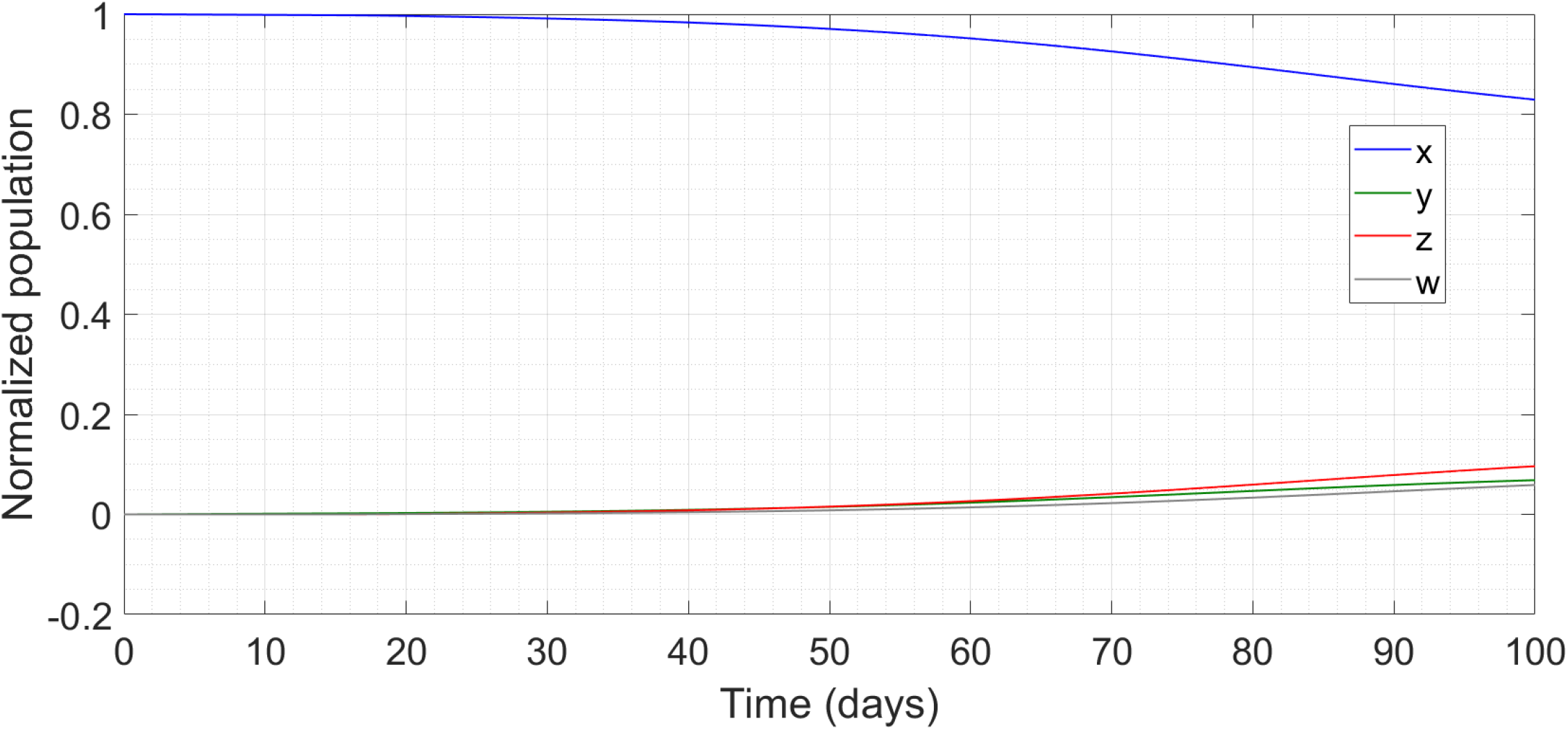
Simulation of the model with reduced transmissibility.

**Figure 4:**
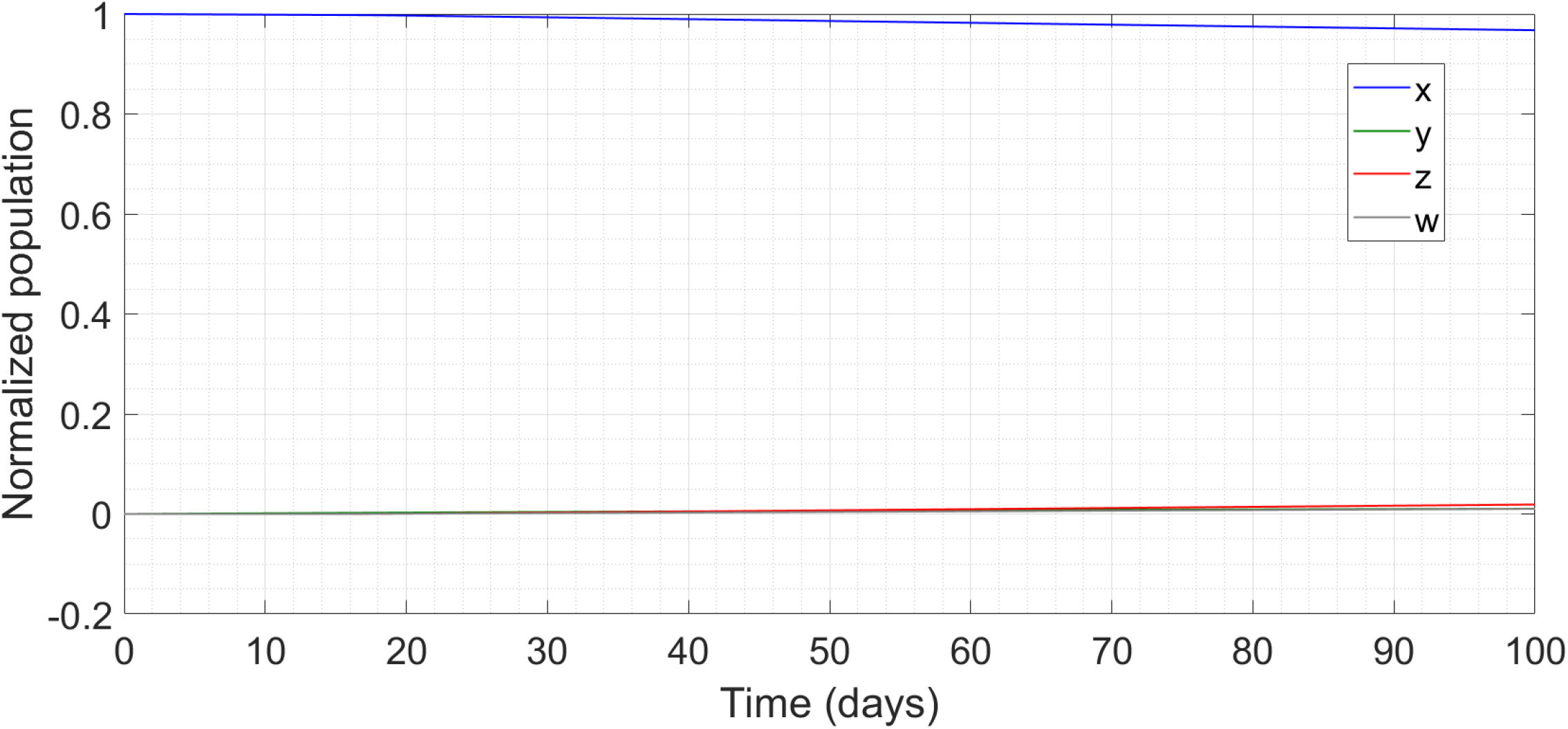
Simulation of the model with reduced latency.

**Figure 5:**
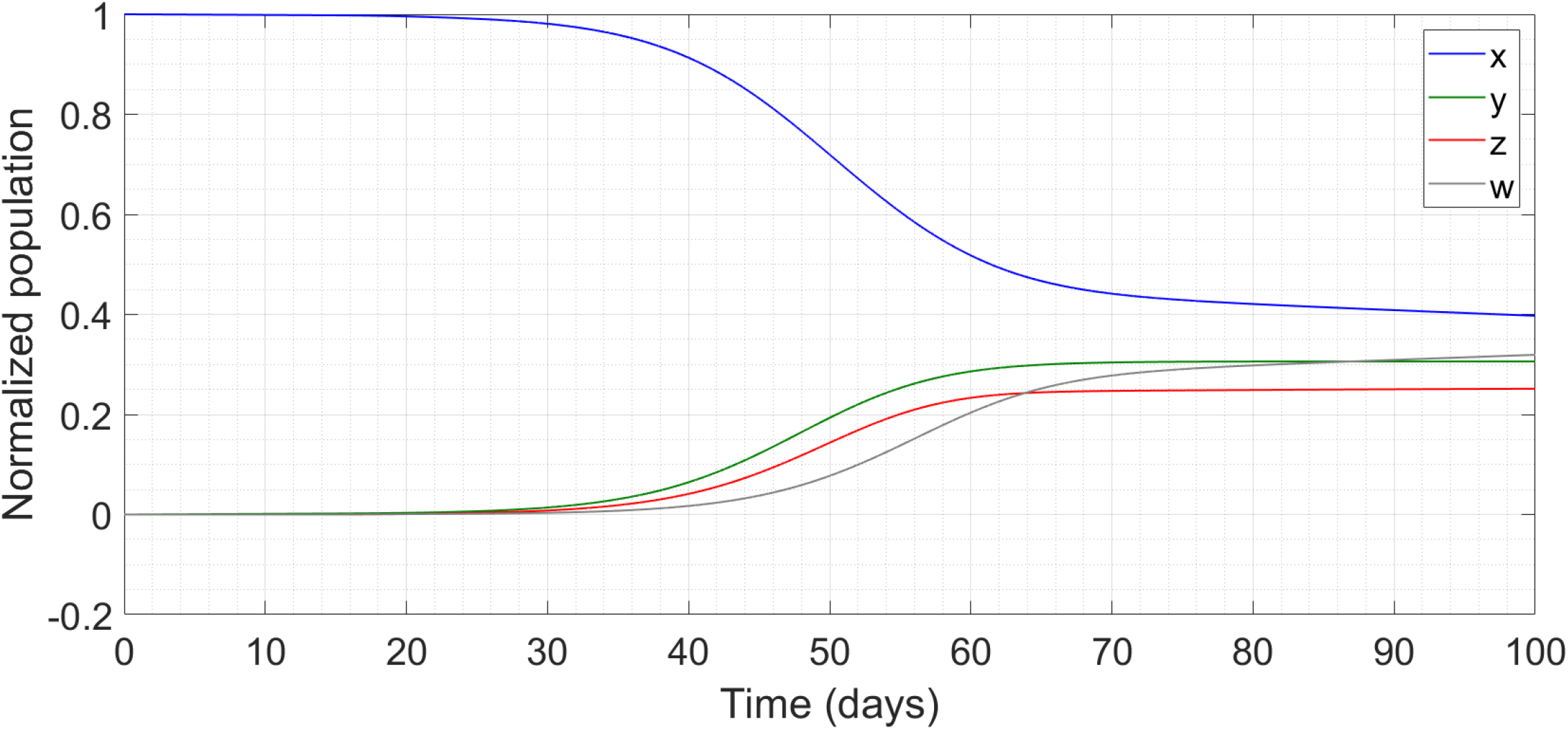
Simulation of the model with reduced fraction of asymptomatic carriers.

A mere factor-of-2 reduction in the latency has made the difference between epidemic and no epidemic.

Finally, another variable which gets discussed in literature is the effect of asymptomatic carriers. Common sense argues that the greater the fraction *μ*_1_ of these carriers, the greater the spread of the disease. In the below plot, we consider the parameter values of Fig. 2 but with *μ*_1_ reduced from 0.18 to 0.10.

There is no drastic change, but the remaining fraction of healthy at large has increased to 40 percent, consistent with our expectations.

So far we have talked about the model in terms of parameter values which are not tailored to any particular region. Now we demonstrate an application to real-world data. We consider the case history of Coronavirus in South Korea, a country which has clearly moved past the peak and is in the declining phase. The day-by-day profile of cases has been obtained from Reference [52]; considering the zeroth day to be the earliest day when there are more than 100 cases (20 February 2020, 104 cases), we find that the rate peaked at around the 10^th^ day when the number of cases was about 3500 and has declined steadily thereafter. The data ends on the 33^rd^ day (yesterday) with the number of cases at 9037. The raw data is given below, as Table 1. An interesting feature of the data is that the growth rate rises to a maximum of about 700/day on days 08-10, then falls rapidly upto day 20 but thereafter becomes almost constant at 100/day for the next 13 days as the cases increase linearly with time.

**Table 1:** Profile of COVID-19 epidemic in Korea.

|  |  |  |  |  |  |  |  |  |  |  |  |
| --- | --- | --- | --- | --- | --- | --- | --- | --- | --- | --- | --- |
| **Day** | **01** | **02** | **03** | **04** | **05** | **06** | **07** | **08** | **09** | **10** | **11** |
| **Cases** | 204 | 346 | 602 | 763 | 977 | 1261 | 1766 | 2337 | 3150 | 3736 | 4212 |
| **Day** | **12** | **13** | **14** | **15** | **16** | **17** | **18** | **19** | **20** | **21** | **22** |
| **Cases** | 4812 | 5328 | 5766 | 6284 | 6767 | 7134 | 7382 | 7513 | 7755 | 7869 | 7979 |
| **Day** | **23** | **24** | **25** | **26** | **27** | **28** | **29** | **30** | **31** | **32** | **33** |
| **Cases** | 8086 | 8162 | 8236 | 8320 | 8413 | 8565 | 8652 | 8799 | 8897 | 8961 | 9037 |

This data has to be fitted to a curve for *w*(*t*), obtained from our model (7). When fitting, the first thing to note is that South Korea has a population of 5,17,00,000 [53], in comparison to which the number of cases is negligible. This is because the bulk of the outbreak occurred in several localized regions which were very effectively cut off from mingling with other, uncontaminated regions. Hence, the initial *x* in this case will not be South Korea’s entire population but an effective value which takes into account the heavily localized character of the affected regions, where mixing of population took place. We find that this effective value naturally comes out of an attempt to fit the known curve. Since *τ*_1_ and *τ*_2_ represent biological properties of the virus, we keep them fixed at 7 and 3 days respectively. South Korea has been extremely proactive at testing, so we set *τ*_3_ = 2. We keep the seeding period at 15 days, assuming linear growth of *y* and *w* during this phase. We choose the growth rate so that *w* becomes equal to 100 at the start of the free evolution and make the ad hoc assumption that at this time there are thrice as many undetected cases as reported ones (this assumption is harmless – changing the number from 3 to 10 makes almost no difference with respect to the trajectory of the disease). Thereafter, we find the following effects of varying parameters :

- To the largest extent, increasing *k*_0_ brings the peak in *w* closer to the start of free evolution of the disease
- The slope of *w* in the post-peak linear regime is governed by *k*_3_ and *μ*_2_
- The initial value of *x* determines the absolute size of *w*
- The parameter *k*_4_ has little effect on the dynamics

We find a good fit to the South Korea data for the following parameter values : *τ*_1_ = 7, *τ*_2_ = 3, *τ*_3_ = 2, *x* (0) = 12900, *k*_0_ = 0.000063, *k*_3_ = 0.0004, *k*_4_ = 0.00004 *μ*_1_ = 0.18 and *μ*_2_ = 0.04. We plot the run upto a time of 60 days, 11 days beyond the available data records.

We can see that the model is not only a good fit to the peak phase of the epidemic but it also correctly predicts the slow yet persistent linear growth in cases after the peak. We further see that the effective *x* (0) is about 13000, implying that approximately 2/3 of the originally susceptible people have been affected. This fraction is similar to the predictions commonly made with respect to the spread of the virus in free society if steps are not taken to reduce social contacts. We again note however, that a fit of the model to data from an entire country is heuristic at best on account of the huge spatial variation of the infection rate, and the model’s assumption of homogeneous mixing among the population. In actual practice, it will be much more useful an exercise to attempt fitting the data to the statistics of a city, district or county, where there is more uniform mixing among the entire population. In such a case, *x* (0) will also not be a completely unknown quantity but will start from a reasonably well-known figure.

On the basis of the above discussion, we posit that the model (7) is a faithful replication of the transmission dynamics of COVID-19. Accordingly, we now use it for predictive purposes, to glimpse results which are as yet in the future and suggest the course of action to be taken accordingly.

## §3 PREDICTIVE RESULTS AND IMPLICATIONS ON POLICY

Considering the benchmark case, we now plot a graph of the derivatives 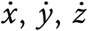 and 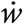 as functions of time. These are basically the daily increments in the populations of various kinds. Recall that *y* is the number of sick people in free society while *w* is the number of positive cases, reported in official records. Hence 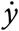 is a measure of the actual strength of the epidemic while 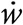 is a measure of the strength as interpreted from available data. The Figure is given below.

We can see that the peak in *y* (green) comes significantly before the peak in *w* (grey). The time interval between the two peaks is approximately 8 days, which corresponds to *τ*_2_ + *τ*_3_. To further bolster this conjecture, we increase *τ*_2_ to 4 (which makes an even more virulent epidemic with 20 percent of *x* remaining after 100 days) and reduce *τ*_3_ to 1. The plot is given below. This time, there is a 5-6 day separation between the two peaks, as we would expect.

Other runs, not shown here, with different values of *k*_0_ and *τ*’s also bring out the truth of the conjecture that the peak in *w* occurs a time approximately *τ*_2_ + *τ*_3_ after the peak in *y*. Thus, the numerical values of the parameters are actually not too important so far as this prediction is concerned. Note that *τ*_1_, technically also a delay term, does not enter here. The lag between actual and reported results is entirely on account of the two delays which are novel to the Coronavirus. Going back to Figure 6, we can see that when *y* has peaked, *w* is still strongly in the ascending region. This implies that, in the initial stages where the numbers of reported cases are increasing strongly, the disease is actually much closer to its peak and spreading much faster than the numbers would indicate. Hence, measures such as lockdowns and enforced social isolation, which reduce the spread of the disease by reducing *k*_0_, need to be implemented as soon as there is a significant growth in the numbers being reported. Waiting for the disease to get “closer to peak” before implementing lockdowns can actually be a sub-optimal policy as that will enforce the curbs after the disease has already spread like wildfire. There is however a brighter side to this picture, which is that when *w* has peaked, *y* is well into the trailing region. Thus, the epidemic is closer to the end that we would otherwise imagine on the basis of a raw interpretation of the data. (As a parenthetical note, this disparity between the actual and reported case histories would have been impossible to obtain from a traditional S-I-R model, which validates our unconventional choice of basic variables.)

**Figure 6:**
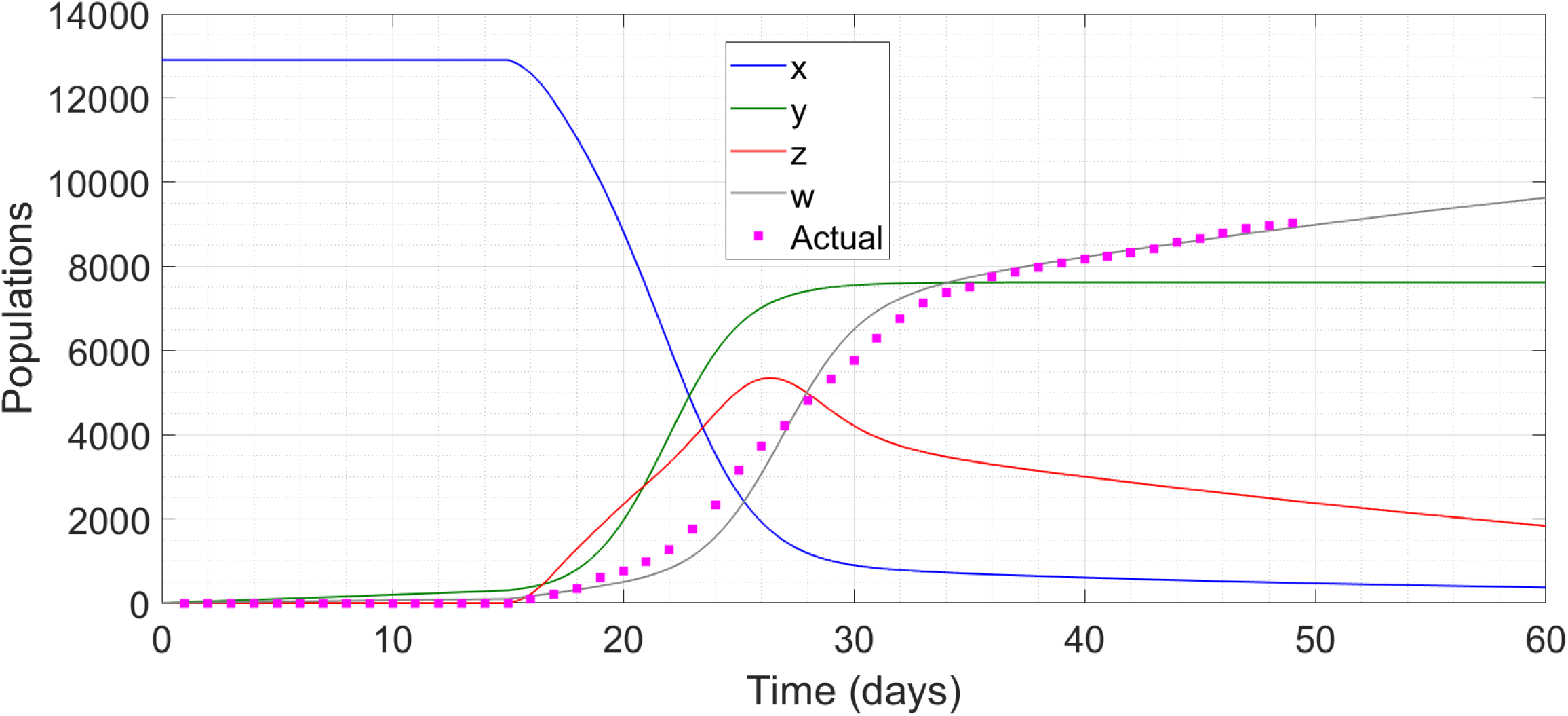
Simulation of the model for Korea and comparison with the actual data set.

We now consider the effect of imposing lockdowns on the spread of the disease. Recall that the degree of social mixing is represented by the parameter *k*_0_. The benchmark results of Figs. 2 and 6 are obtained for *k*_0_ = 0.0008, and we have already seen that reducing this parameter improves the time history of spread of the disease. We assume a value of *k*_0_ = 0.0005 during a lockdown phase. If this figure is maintained throughout the 100-day period then there is almost no spread of the epidemic with 98 percent of healthy at large population remaining at the end of the period. However, an indefinite lockdown from start to stop is unrealistic and impossible to implement. A practical lockdown must be of a limited duration. Accordingly, we consider the effects of a 15-day lockdown being implemented at various stages in the progress of the disease.

Recall that in our model the free evolution of the disease starts after 15 days, and that in the base case (no lockdown) the peak of *y* occurs at 45 days, the peak of *w* occurs at 53 days, and the fraction of *x* remaining at 100 days is 31 percent. For the first run, we implement a very early lockdown from *t* = 20 to *t* = 35.

This is an ineffective move – the disease remains quiescent during the lockdown but explodes after the restrictions are lifted. A similar phenomenon also happens for a range of early lockdowns, thus nullifying their effect completely. This runs contrary to the perception that locking down at the earliest signs of spread is the strategy to adopt – it is good if the lockdown can be indefinite but inutile when there are external constraints on its duration. Similarly, a lockdown at too late a stage (after the disease has peaked) is also useless, for obvious reasons. We find that the optimal time of imposing the lockdown is such that the centre of the lockdown period corresponds to the peak in *y* in the absence of the lockdown. For the next run, we impose lockdown from *t* = 37 to *t* = 52, straddling the *y*-peak at *t* = 44. The results are presented below.

This well-timed lockdown cripples the flow of the disease before it approaches its peak. Moreover, the disease does not restart its virulent spread after the curbs are eased. The remaining fraction of *x* at the end of the 100-day period turns out to be 53 percent. Instead, if we impose the 15-day lockdown with its centre coinciding with the (no lockdown) peak in *w*, i.e. from *t* = 45 to *t* = 60, then too the flow of the disease is choked but the remaining fraction of *x* reduces to 45 percent, which is a nearly 1/5 drop from the previous case. In the next plot, we vary the starting point of the 15-day lockdown from the 20^th^ day to the 55^th^ day and plot the remaining percentage of *x* at the end of the 100-day period. We can see a clear maximum in this percentage when the lockdown is timed to straddle and hence stamp out the peak in *y*; the percentage decreases rapidly on both sides of the maximum.

Another feature of the bottom panel of Fig. 10 is extremely important. We see from the grey curve that the **rate of detected cases d*w*/d*t* keeps increasing even after the lockdown has been imposed**. Indeed, in a qualitative sense, 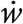 in the presence of the lockdown proceeds smoothly to a peak and then tails off, just as it would have in the absence of the lockdown (there is a spike at the instant the lockdown is lifted but that is probably a numerical artefact and unlikely to manifest in practice). This might mislead one to believe that the lockdown has been ineffective. However, the actual decrease, and a very significant one at that, has taken place in a quantity (*y*) which cannot be measured directly. The height of the peak in 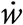 with the lockdown in place (about 0.007) has also reduced drastically relative to that without the lockdown (Fig. 7, height about 0.019) – once again, a comparison with an unmeasurable quantity. Thus we see that the lockdown serves its purpose even if this is not apparent from the limited data (detected case trajectory) at our disposal. This is very likely the explanation behind the apparent futility of the lockdowns imposed in Italy and Spain, where the numbers of cases continue to rise aggressively even after the most drastic social isolation measures have been implemented. We shudder to think what these numbers would have been if the lockdowns had not been imposed when they were. We also note that the apparent lack of qualitative change after a lockdown can only be found from our delayed model and not from a real-time S-I-R model for example Reference [42].

**Figure 7:**
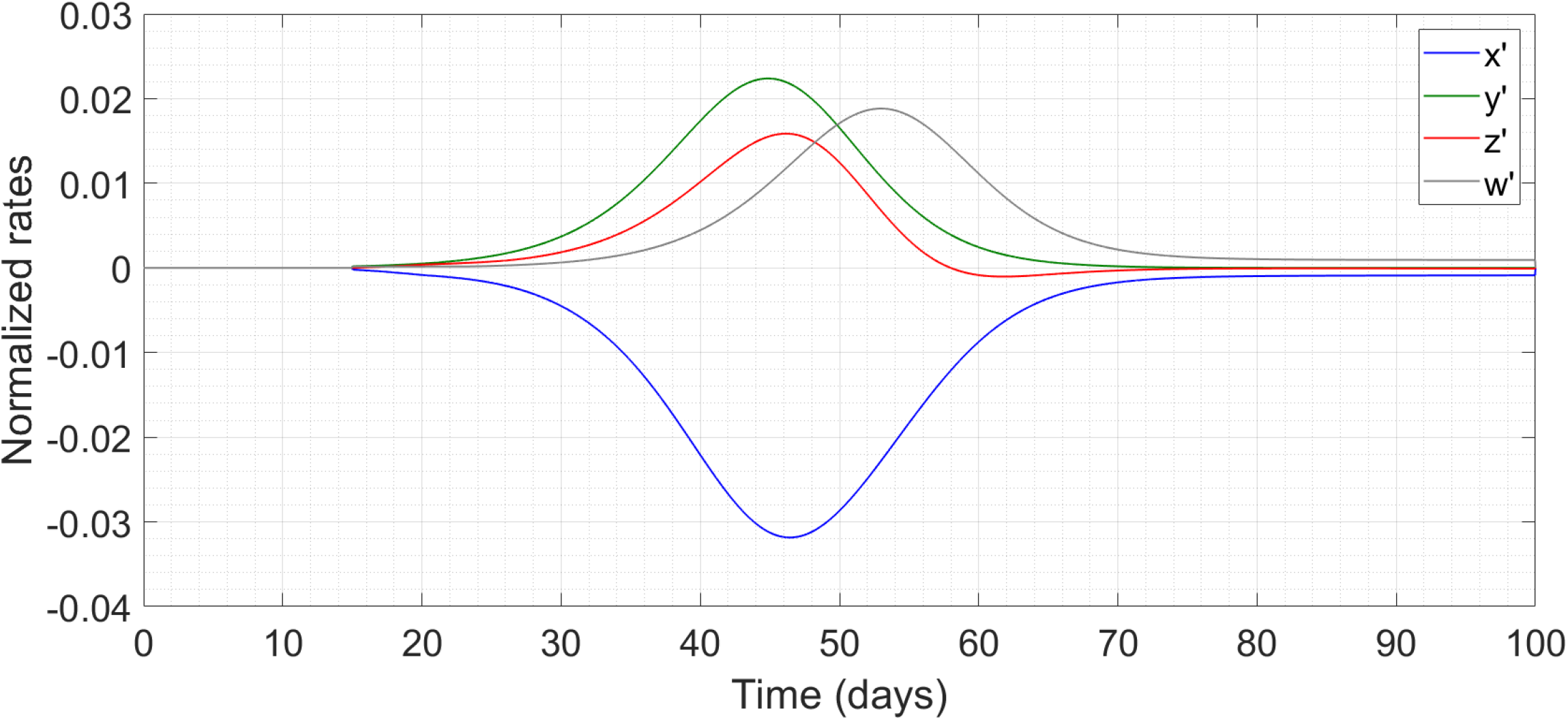
Simulation of the benchmark model showing the growth rates.

**Figure 8:**
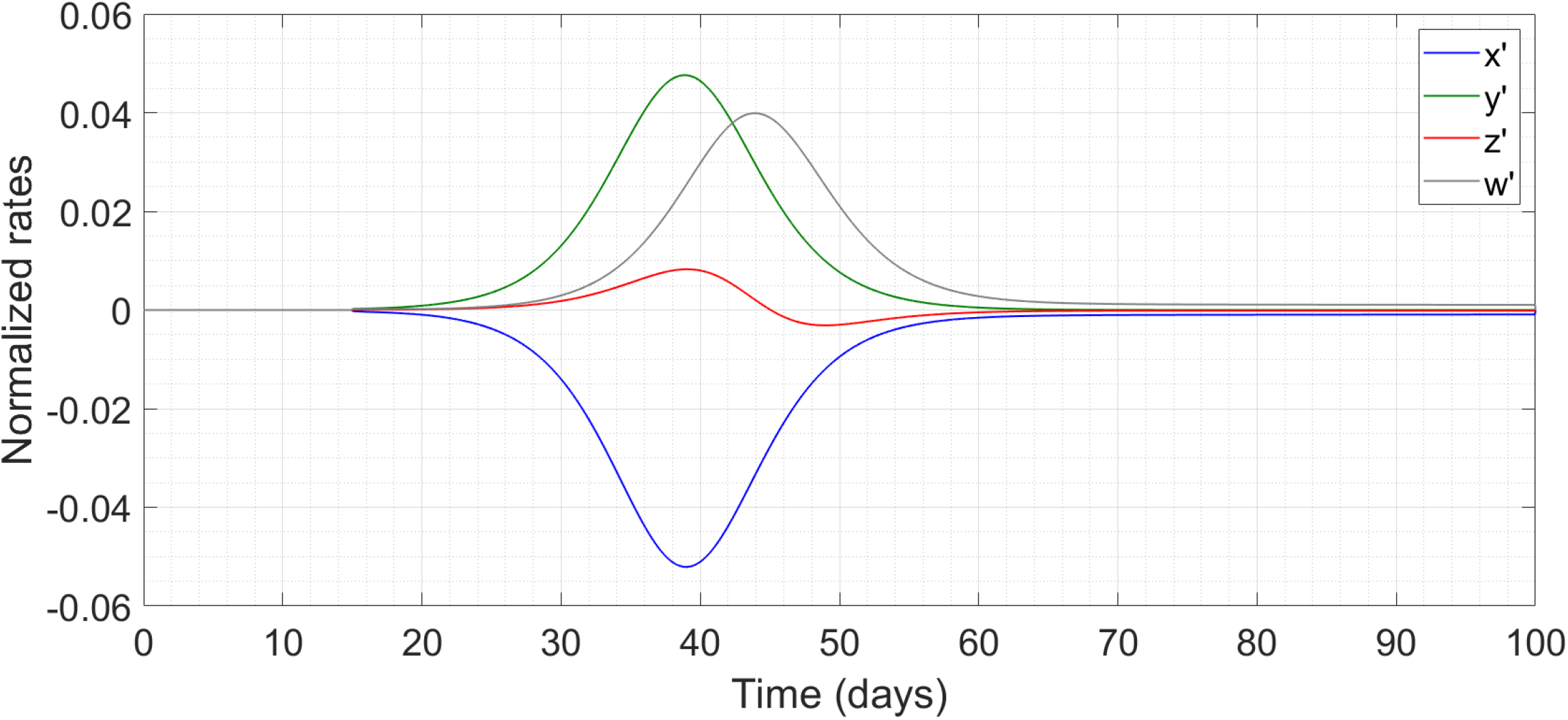
Simulation of the model with altered delays.

**Figure 9:**
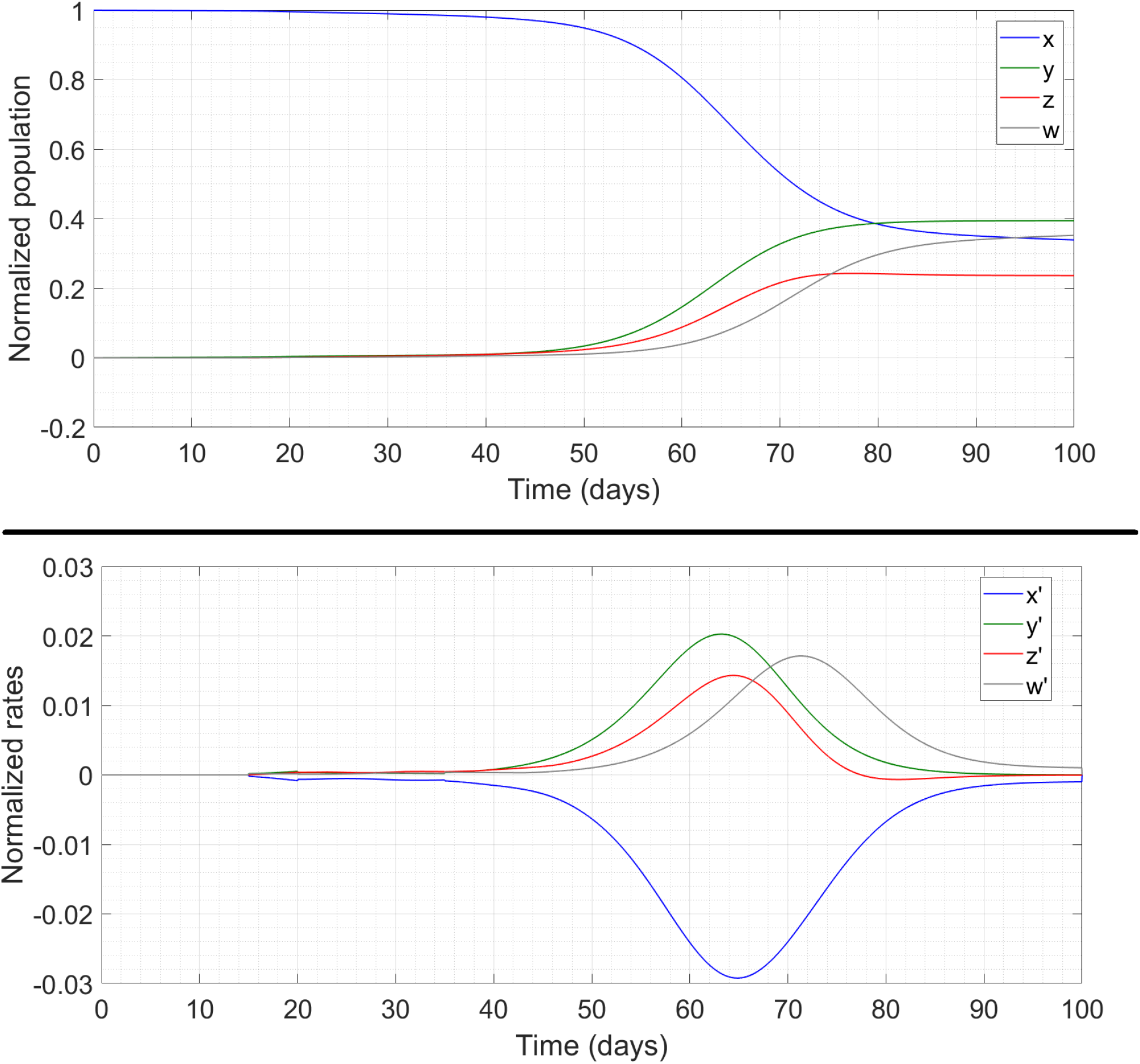
Simulation of the model with early lockdown. The left panel shows the populations while the right panel shows the rates.

**Figure 10:**
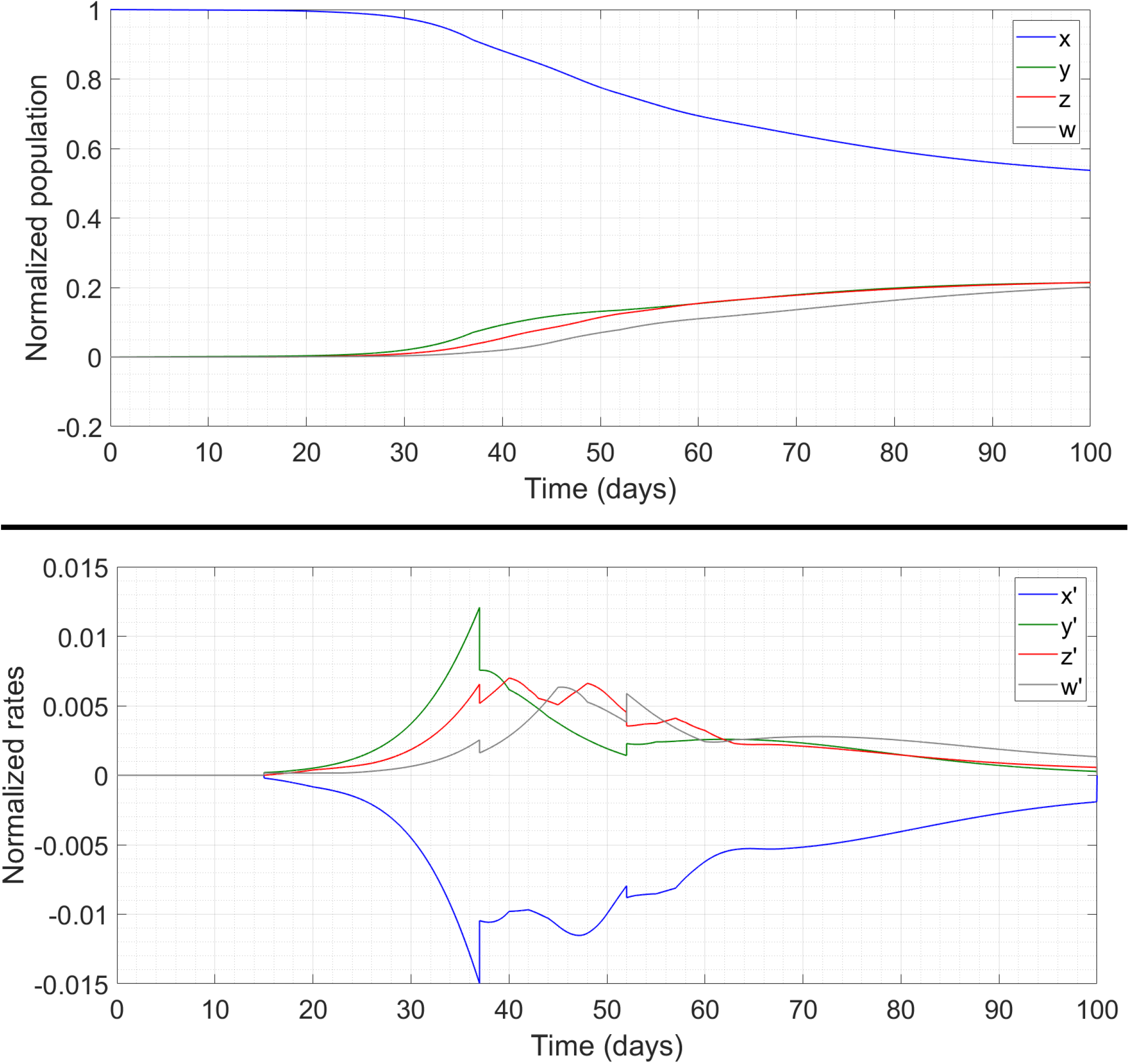
Simulation of the model with a well-timed lockdown. The top panel shows the populations while the bottom panel shows the rates.

We now discuss how our results can be used by the regional authorities of a city, district or county. We consider a region which is still approaching the peak and has implemented zero or partial social distancing measures. The biological parameters *τ*_1_, *τ*_2_ and *μ*_1_ will remain fixed, at the values used here or at values which the authorities obtain from their (likely more comprehensive) data sets. The initial value *x* (0) will be some fraction of the total population of the region (accounting for people who are resistant, people who are very seldom outdoors etc). Authorities can use their testing records to find approximate values of *τ*_3_, *k*_3_, *k*_4_ and *μ*_2_. Thereafter, the extant data for *w* (*t*) can be fitted to the model to determine a basic *k*_0_, and further refine the estimates of the other parameters. Given these, the authorities can now find the location of the expected peak in *y*, and impose a limited-duration lockdown centred round that peak. Figure 11 indicates that there **is** some margin of error here – the remaining percentage stays above 50 if the lockdown is imposed any time between the 34^th^ and 42^nd^ day. This is a considerably large window period which can account for inaccuracies in parameter estimation, disparities between model and reality etc.

**Figure 11:**
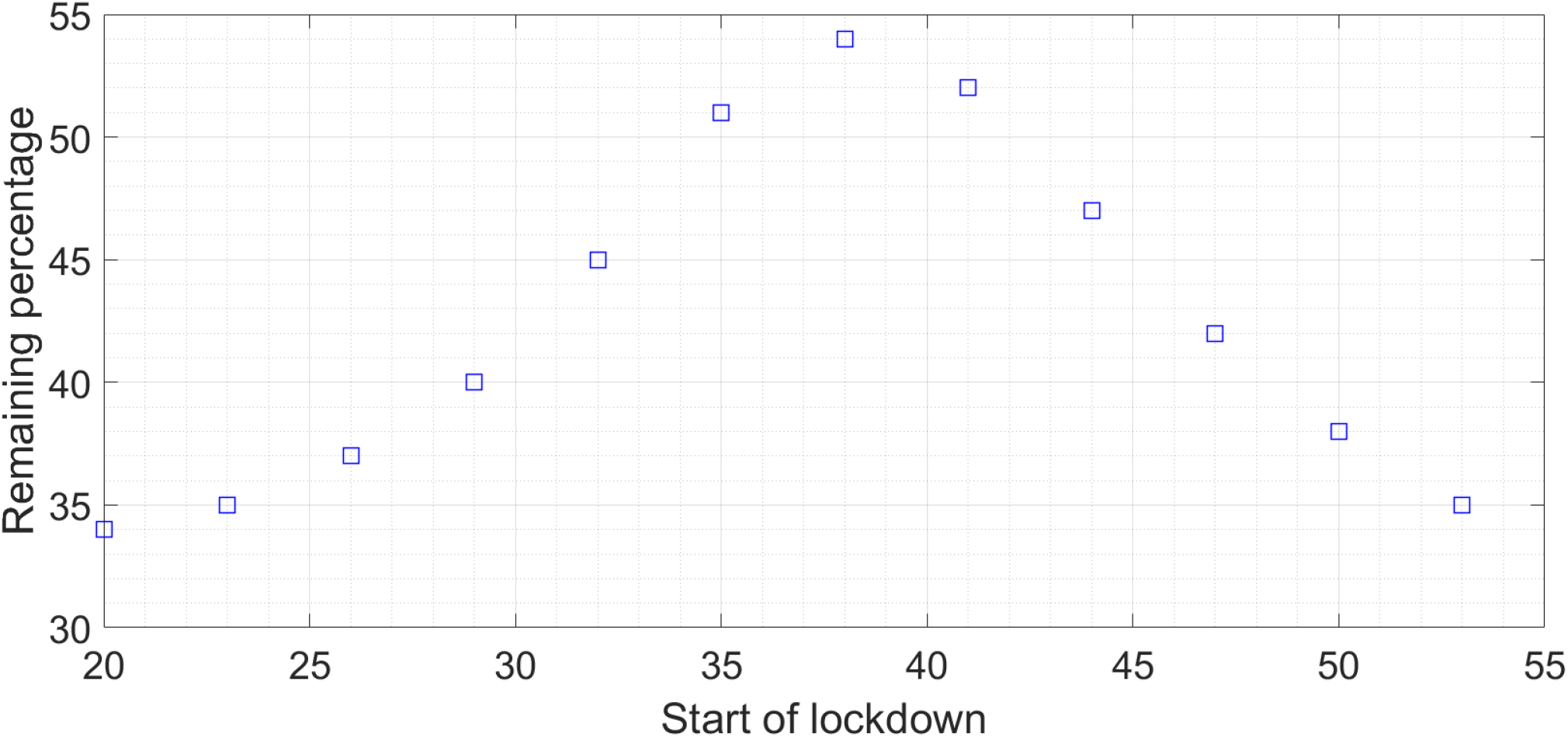
Plot of the remaining percentage of × if the 15-day lockdown is started at various times in the progression of the disease.

## §4 CONCLUSIONS AND FUTURE DIRECTIONS

In this work we have constructed a novel lumped-parameter mathematical model of COVID-19 which incorporates the effects of the latency period between transmissibility and symptomaticity as well as of the delay in testing on account of the limited testing facilities currently available. We have used this model to predict the effects of drastic but short-term social distancing measures on the spread of the disease. Our principal findings are that :

- The delays cause the peak of the actual spread of the epidemic to come a few days before the reported peak.
- The most effective interval for imposing a limited-duration lockdown is one which straddles the peak of the spread of the disease in society in the absence of the lockdown.
- After imposing a lockdown, the trajectory of the number of reported cases may show no outwardly visible change even if the lockdown has been well-timed and has significantly slowed the spread of the disease.
- Extensive planning to allocate the resources for management of cases, measures like quarantine and social distancing and simultaneously building capacity for innovative approaches to arrange for testing kits should be executed in the initial stages of the epidemic. The required capacities of quarantine and treatment facilities can be estimated early on from the predicted time traces of *z* (*t*).

For impactful epidemic management, societies need to model their strategies, develop an advanced surveillance system for early detection of cases, facilitate implementation of preventive interventions, and enable effective coordination of healthcare professionals, public health authorities, policy makers and government. Subsequent work will have to focus on extending and refining our model. For example, one can take into account the effects of errors in testing and intentional or unintentional violations of quarantine. According to the Institute of Medicine (IoM) categories, drivers of pathogen emergence like human demographics and behaviour, economic development and land use, international travel and commerce, changing ecosystems and human susceptibility should also be taken into consideration. One can also integrate our lumped-parameter model with more detailed analyses which account for heterogeneous spatial mixing and stochasticity in contracting the disease. Such steps will increase even more the predictive power of the model, and further help regional authorities in timing their lockdowns, and estimating the required capacities of quarantine and treatment facilities.

## Data Availability

All data is available from websites mentioned in the References.

## Notes

### Competing Interest Statement

The authors have declared no competing interest.

### Funding Statement

No external funding was received for this study.

## REFERENCES

[1] C Huang et. al., “Clinical features of patients infected with novel coronavirus in Wuhan, China,” The Lancet 395 (10223), 497–506 (2020)

[2] So rapidly are the numbers changing that during revision of this Article on 29 March 2020, the total number of cases has exceeded 7,00,000 worldwide, and the countries with maximal numbers of cases are USA, Italy and only then China (assuming that all reported data sets are true and accurate).

[3] JF Chan et. al., “Interspecies transmission and emergence of novel viruses : lessons from bats and birds,” Trends in Microbiology 21 (10), 544–555 (2013)

[4] Y Chen, Q Liu and D Guo, “Emerging coronaviruses : genome structure, replication, and pathogenesis,” Journal of Medical Virology 92 (4), 418–423 (2020)

[5] JF Chan et. al., “Genomic characterization of the 2019 novel human-pathogenic coronavirus isolated from a patient with atypical pneumonia after visiting Wuhan,” Emerging Microbes and Infections 9 (1), 221–236 (2020)

[6] R Lu et. al., “Genomic characterisation and epidemiology of 2019 novel coronavirus : implications for virus origins and receptor binding,” The Lancet (2020)

[7] P Zhou et. al., “A pneumonia outbreak associated with a new coronavirus of probable bat origin,” Nature (2020)

[8] X Li, Y Song, G Wong and J Cui, “Bat origin of a new human coronavirus: there and back again,” Science China Life Sciences 63 (3), 461–462 (2020)

[9] W Li et. al., “Bats are natural reservoirs of SARS-like coronaviruses,” Science 310 (5748), 676–679 (2005)

[10] D Paraskevis et. al., “Full-genome evolutionary analysis of the novel corona virus (2019-nCoV) rejects the hypothesis of emergence as a result of a recent recombination event,” Infection, Genetics and Evolution 79, 104212 (2020)

[11] LE Gralinski and VD Menachery, “Return of the Coronavirus : 2019-nCoV,” Viruses 12 (2), 135 (2020)

[12] MC Wong, SJJ Cregeen, NJ Ajami and JF Petrosino, “Evidence of recombination in coronaviruses implicating pangolin origins of nCoV-2019,” BioRxiv (2020)

[13] K Xiao et. al., “Isolation and characterization of 2019-nCoV-like coronavirus from Malayan pangolins,” BioRxiv (2020)

[14] X Tang et. al., “On the Origin and continuing evolution of SARS-CoV-2,” National Science Review, waa036

[15] P Zhou et. al., “Discovery of a novel coronavirus associated with the recent pneumonia outbreak in humans and its potential bat origin,” BioRxiv (2020)

[16] SA Lauer et. al., “The Incubation period of coronavirus disease 2019 (COVID-19) from publicly reported confirmed cases : estimation and application,” Annals of Internal Medicine (2020)

[17] Q Li et. al., “Early Transmission Dynamics in Wuhan, China, of Novel Coronavirus-Infected Pneumonia,” New England Journal of Medicine (2020)

[18] CT Bauch, JO Lloyd-Smith, MP Coffee and AP Galvani, “Dynamically modeling SARS and other newly emerging respiratory illnesses : past, present, and future,” Epidemiology 16 (6), 791–801 (2005)

[19] Y Liu, AA Gayle, A Wilder-Smith and J Rocklov, “The Reproductive number of COVID-19 is higher compared to SARS coronavirus,” Journal of Travel Medicine (2020)

[20] S Knobler et. al. (Eds.), “Learning from SARS: Preparing for the Next Disease Outbreak,” Workshop Summary, National Academies Press, Washington DC, USA (2004)

[21] J Wu et. al., “Risk factors for SARS among persons without known contact with SARS patients in Beijing, China,” Emerging Infectious Diseases 10 (2), 210–216 (2004)

[22] MY Yen et. al., “Taiwan’s traffic control bundle and the elimination of nosocomial severe acute respiratory syndrome among healthcare workers,” Journal of Hospital Infection 77 (4), 332–337 (2011)

[23] W Kermack and AG McKendrick, “A Contribution to the mathematical theory of epidemics,” Proceedings of the Royal Society A 115 (772), 700–721 (1927)

[24] S Towers, KV Geisse, Y Zheng and Z Feng, “Antiviral treatment for pandemic influenza : assessing potential repercussions using a seasonally forced SIR model,” Journal of Theoretical Biology 289, 259–268 (2011)

[25] A Pandey, A Mubayi and J Medlock, “Comparing vector-host and SIR models for dengue transmission,” Mathematical Biosciences 246 (2), 252–259 (2013)

[26] A Rachah and DFM Torres, “Predicting and controlling the Ebola infection,” Mathematical Methods in Applied Sciences 40 (17), 6155–6164 (2017)

[27] E Beretta and Y Takeuchi, “Global stability of an SIR epidemic model with time delays,” Journal of Mathematical Biology 33 (3), 250–260 (1995)

[28] CC McCluskey, “Complete global stability for an SIR epidemic model with delay - distributed or discrete,” Nonlinear Analysis : Real World Applications 11 (1), 55–59 (2010)

[29] AJ Kucharski et. al., “Early dynamics of transmission and control of COVID-19 : a mathematical modeling study,” The Lancet Infectious Diseases (2020)

[30] Y Fang and Y Nie, “Transmission dynamics of the COVID-19 and effectiveness of government interventions : a data-driven analysis,” Journal of Medical Virology (2020)

[31] JF Rabajante, “Insights from early mathematical models of 2019 n-CoV acute rspiratory disease (COVID-19) dynamics,” Arxiv article 2002.05296

[32] L Peng et. al., “Epidemic analysis of COVID-19 in China by dynamical modeling,” Arxiv article 2002.06563

[33] TM Chen et. al., “A Mathematical model for simulating the phase-based transmissibility of novel coronavirus,” Infectious Diseases of Poverty (2020)

[34] RA Neher et. al., “Potential impact of seasonal forcing on SARS-CoV-2 pandemic,” Swiss Medical Weekly 150, w20224 (2020)

[35] K Mizumoto, K Kagaya and G Chowell, “Early epidemiological assessment of the transmission potential and virulence of coronavirus disease 2019 (COVID-19) in Wuhan city, China in January-February 2020,” Medrxiv preprint (2020)

[36] K Mizumoto and G Chowell, “Transmission potential of the novel coronavirus (COVID-19) on board the Diamond Princess cruise ship 2020,” Infectious Disease Modeling 5, 264–270 (2020)

[37] NM Ferguson et. al. “Impact of non-pharmaceutical interventions (NPI) to reduce COVID-19 mortality and healthcare demand,” electronically available at https://www.imperial.ac.uk/media/imperial-college/medicine/sph/ide/gida-fellowships/Imperial-College-COVID19-NPI-modelling-16-03-2020.pdf (2020)

[38] NM Ferguson et. al., “Strategies for containing an emerging influenza pandemic in South-East Asia,” Nature 437 (7056), 209–214 (2005)

[39] JR Koo et. al., “Interventions to mitigate early spread of SARS-CoV-2 in Singarpore : a modeling study,” The Lancet Infectious Diseases (2020)

[40] DL Chao, ME Halloran, VJ Obenchain and IM Longini, “FLUTE : A Publicly available stochastic epidemic influenza simulation model,” PLoS Computational Biology 6 (1), e1000656 (2010)

[41] S Maslov and N Goldenfeld, “Window of opportunity for mitigation to prevent overflow of ICU facilities in Chicago by COVID-19,” Arxiv article 2003.09564

[42] R Singh and R Adhikari, “Age-structured impact of social distancing on the COVID-19 epidemic in India,” Arxiv article 2003.12055

[43] KK To et. al., “Temporal profiles of viral load in posterior oropharyngeal saliva samples and serum antibody responses during infection by SARS-CoV-2 : an observational cohort study,” The Lancet Infectious Diseases (2020)

[44] https://www.cdc.gov/coronavirus/2019-ncov/cases-updates/testing-in-us.html

[45] P Nouvellet et. al., “The Role of rapid diagnostics in managing Ebola epidemics,” Nature 528 (7580), S109–S116 (2015)

[46] E Vergu, H Buson and P Ezanno, “Impact of the infection period distribution on the epidemic spread in a metapopulation model,” PLoS ONE 5 (2), e9371 (2010)

[47] G Onder, G Rezza and S Brusafero, “Case fatality rate and characteristics of patients dying in relation to COVID-19 in Italy,” Journal of the American Medical Association (2020)

[48] LF Shampine and S Thompson, “Solving DDEs in Matlab,” Applied Numerical Mathematics 37 (4), 441–448 (2001)

[49] M Davidow, B Shayak and RH Rand, “Analysis of a remarkable singularity in a nonlinear DDE,” Nonlinear Dynamics 90 (1), 317–323 (2017)

[50] K Mizumoto, K Kagaya, A Zarebski and G Chowell, “Estimating the asymptomatic proportion of coronavirus disease 2019 (COVID-19) cases on board the Diamond Princess Cruise Ship, Yokohama, Japan, 2020,” Eurosurveillance 25 (10), 1–10 (2020)

[51] S Cauchemez et. al., “Middle East respiratory syndrome coronavirus : quantification of the extent of the epidemic, surveillance biases and transmissibility,” The Lancet Infectious Diseases 14 (1), 50–56 (2014)

[52] https://www.bing.com/covid

[53] https://en.wikipedia.org/wiki/South_Korea

